# Red flags to screen for tumors in patients with neck pain: a scoping review protocol

**DOI:** 10.1101/2024.05.22.24307749

**Authors:** Beatrice Occhetto, Martina Ballesio, Firas Mourad, Marco Trucco, Filippo Maselli, Alessandro Chiarotto, Daniel Feller

## Abstract

**Background:** People with malignant (primary or metastatic) or benign tumors may present in clinical practice with neck pain, with or without other symptoms (e.g., radicular pain or headache). When not presenting as an emergency, neck pain is most often assessed by primary care clinicians such as general practitioners, physiotherapists, chiropractors or osteopaths. Therefore, primary care clinicians must be able to screen for tumors when evaluating patients with neck pain.

Despite clinical practice guidelines providing recommendations to triage serious conditions presenting as neck pain, there is a paucity of overviews on red flags for tumors in patients presenting with neck pain in primary care settings. The present scoping review aims to comprehensively map the existing literature on red flags for tumors in patients presenting with neck pain in primary care settings. Furthermore, we will aim to identify gaps in the literature to direct future research in this area.

**Methods:** We will search MEDLINE (via PubMed), Embase, CINHAL, and Scopus. In addition, we will use Web of Science to implement backward and forward citation tracking strategies. We will consider any primary study design written in English or Italian. No time or geographical restrictions will be applied to the search. Studies with a focus on the diagnostic pathway, considering patients of any age and gender with a diagnosis of tumor and a primary complaint of neck pain will be eligible for inclusion. Only studies conducted in primary care settings will be considered. Two authors will independently perform the study selection and data extraction phases. Results from the scoping review will be summarized descriptively through tables and diagrams. As a scoping review, we will highlight any gaps in the existing literature regarding our research questions.

## BACKGROUND

Neck pain is a highly prevalent condition that leads to considerable pain, disability, and economic cost. A systematic analysis of the Global Burden of Disease Study 2021 states that in 2020 neck pain affected 203 million people worldwide^(1)^. The majority of people complaining of musculoskeletal neck disorders are classified as “non-specific”, as no identifiable underlying diseases or abnormal anatomical structure may be linked to the patient’s symptoms^(1)^. Rarely, neck pain may be the early manifestation of more serious conditions, such as cervical myelopathy, cervical ligamentous instability, fracture, tumor, vascular insufficiency, systemic diseases or other specific causes of spinal or non-spinal origins. These patients may seek help and require support mainly in a primary care pathway^(2)^. Therefore, early triage is a mainstay for primary care clinicians^(3)^.

Primary tumors of the cervical spine are relatively rare, with a global incidence rate ranging from 2.5 to 8.5 cases per 100,000 people per year^(4)^. Although the incidence of primary cervical spinal tumors is low, mortality and disability rates are high, especially among young patients^(4)^. Spinal malignancy, also known as metastatic bone disease, refers to metastases that have spread specifically into the spinal bones. These metastases are common in various primary cancers, such as breast, prostate, lung, kidney, and thyroid, affecting the cervical spine 10% of the time^(5)^. Benign tumors of the spine, tumors of neural tissue, calcifying-pseudo neoplasm of the spine, meningioma, malignant tumors, metastases of epithelial tumors, and tumors of hematopoietic tissue are also reported to be associated with neck pain^(6)^. Red flags are specific indicators derived from a patient’s medical history and physical examination (e.g., history of cancer, nocturnal pain, visual disturbances) signaling a heightened risk of serious pathologies, such as tumors^(3)^. Recognizing red flags can aid clinicians in the early detection of serious pathology and prompt additional assessments, resulting in better outcomes. ^(7,8)^ However, unnecessary referrals and diagnostic testing need to be balanced against the risk of missing a diagnosis^(9)^. Despite the existing literature providing guidelines for the differential diagnosis of neck pain^(2,6)^, there is a lack of overviews on red flags for tumors in patients presenting with neck pain in primary care settings.

Therefore, the objective of this scoping review is to systematically map and summarize the current literature to identify studies that reported on red flags for tumors in patients presenting with neck pain and associated symptoms (e.g. radicular pain, headache) in primary care settings, to identify gaps in the evidence base and to direct future research in this area. In particular, we will focus on the diagnostic pathway, with attention to the following aspects:

- the type of primary care clinician (e.g., GPs, physiotherapists, chiropractors);
- the pain characteristics (e.g., intensity and localization);
- the prevalence of associated signs and symptoms (e.g., neurological impairments);
- the type of tumor (e.g., primary or metastasis, malignant or benign tumors);
- the demographic characteristics (e.g., gender and age);
- the prevalence of risk factors (e.g., history of tumor);
- other relevant clinical predictors (e.g., comorbidities).

## METHODS

The present scoping review will be conducted in accordance with the Joanna Briggs Institute (JBI) methodology for scoping reviews^(10)^. The Preferred Reporting Items for Systematic Reviews and Meta-Analyses extension for Scoping Reviews (PRISMA-ScR) checklist will be used for reporting the manuscript^(10)^.

### Inclusion criteria

Articles will be eligible for inclusion if they meet the following population, concept, and context (PCC) criteria:

- <u>Population</u>: patients of any age and gender presenting to a primary care clinician with neck pain as the primary complaint and with a final diagnosis of tumor. The diagnosis of tumor can encompass both malignant tumors (primary or metastatic) and benign ones. Patients presenting associated symptoms in addition to neck pain (e.g., radicular pain and headache) will be also included;
- <u>Concept</u>: this review will consider studies that explored and reported on the diagnostic pathway for tumors among the aforementioned population;
- <u>Context</u>: this review will consider studies conducted in primary care settings.

### Sources

This scoping review will include any primary study design written in English or Italian. No time or geographical restrictions will be applied. We will include studies in which at least 80% of the population meets the inclusion criteria or the study provides a separate analysis for patients who meet our inclusion criteria.

### Exclusion criteria

Studies that do not meet the above-stated PCC criteria will be excluded.

### Search strategy

We will search the following databases: MEDLINE (via PubMed), Embase, CINHAL, and Scopus from inception up to 22/05/2024. In addition, we will use Web of Science to implement backward and forward citation tracking strategies.

(“Neck Pain”[MeSH Terms] OR “neck pain*”[Title/Abstract] OR “torticollis”[MeSH Terms] OR “torticollis”[Title/Abstract] OR “cervicalgia*”[Title/Abstract] OR “neck ache*”[Title/Abstract] OR “neckache*”[Title/Abstract] OR “cervicodynia*”[Title/Abstract] OR “cervical pain*”[Title/Abstract] OR “neck injur*”[Title/Abstract] OR “neck disorder*”[Title/Abstract] OR “Wryneck”[Title/Abstract] OR “cervical radiculopathy”[Title/Abstract] OR”cervical disorder*”[Title/Abstract] OR “cervical injur*”[Title/Abstract]) AND (“Neoplasms”[MeSH Terms] OR “neoplasm*”[Title/Abstract] OR “tumor*”[Title/Abstract] OR “cancer*”[Title/Abstract] OR “neoplasia*”[Title/Abstract] OR “malignanc*”[Title/Abstract])

### Study selection

Two reviewers will independently perform the study selection process, firstly by title/abstract and secondly by full-text. Any disagreement will be resolved by consensus or by the decision of a third author. We will use the online electronic systematic review software package (Rayyan QCRI) to organize and track the selection process^(11).^ Reasons for excluding any full-text source of evidence will be recorded and reported.

### Data extraction

The data extraction process will be conducted independently by two reviewers using a standardized form. Any discrepancies will be resolved by a consensus between the two authors and, if necessary, by a third author’s decision. We will aim to extract the following information from each study:

- first author, publication year and country;
- study design;
- type of care clinician (e.g., GPs, physiotherapists, chiropractors);
- pain characteristics (e.g., intensity and localization);
- prevalence of associated signs and symptoms (e.g., neurological signs);
- type of tumor (e.g., malignant or benign tumors);
- demographic characteristics of the included patients (e.g., gender and age);
- prevalence of risk factors (e.g., history of tumor);
- other relevant clinical predictors (e.g., comorbidities).

The process of charting in scoping reviews is iterative. If additional relevant items emerge during full-text analysis, we will extract more data. Any modifications to the data extraction phase will be mentioned in the full manuscript.

### Data synthesis

Results from the scoping review will be summarized descriptively through tables and figures. More specifically, we will narratively summarize and synthesize data relating to the: type of primary care clinician, pain characteristics, prevalence of associated signs and symptoms, type of tumor, demographic characteristics of the included patients, prevalence of risk factors, and other relevant clinical predictors. As a scoping review, we will highlight any gaps in the existing literature regarding our research questions.

## Data Availability

All data produced in the present study are available upon reasonable request to the authors

